# Genetic risk of obesity as a modifier of associations between neighbourhood environment and body mass index: an observational study of 335,046 UK Biobank participants

**DOI:** 10.1101/19004671

**Authors:** Kate E Mason, Luigi Palla, Neil Pearce, Jody Phelan, Steven Cummins

## Abstract

**Background:** There is growing recognition that recent global increases in obesity are the product of a complex interplay between genetic and environmental factors. However, in gene-environment studies of obesity, ‘environment’ usually refers to individual behavioural factors that influence energy balance, while more upstream environmental factors are overlooked. We examined gene-environment interactions between genetic risk of obesity and two neighbourhood characteristics likely to be associated with obesity (proximity to takeaway/fast-food outlets and availability of physical activity facilities).

**Methods:** We used data from 335,046 adults aged 40-70 in the UK Biobank cohort to conduct a population-based cross-sectional study of interactions between neighbourhood characteristics and genetic risk of obesity, in relation to BMI. Proximity to a fast-food outlet was defined as distance from home address to nearest takeaway/fast-food outlet, and availability of physical activity facilities as the number of formal physical activity facilities within one kilometre of home address. Genetic risk of obesity was operationalised by 91-SNP and 69-SNP weighted genetic risk scores, and by six individual SNPs considered separately. Multivariable, mixed effects models with product terms for the gene-environment interactions were estimated.

**Results:** After accounting for likely confounding, the association between proximity to takeaway/fast-food outlets and BMI was stronger among those at increased genetic risk of obesity, with evidence of an interaction with polygenic risk scores (P=0.018 and P=0.028 for 69-SNP and 91-SNP scores, respectively) and in particular with a SNP linked to *MC4R* (P=0.009), a gene known to regulate food intake. We found very little evidence of a gene-environment interaction for availability of physical activity facilities.

**Conclusions:** Individuals at an increased genetic risk of obesity may be more sensitive to exposure to the local fast-food environment. Ensuring that neighbourhood residential environments are designed to promote a healthy weight may be particularly important for those with greater genetic susceptibility to obesity.

## BACKGROUND

Obesity has a heritable component,[1] but the rapid rise in global obesity prevalence suggests an important role for environmental influences.[2] However, individuals may have differing physiological or behavioural responses to the increasingly ‘obesogenic’ environment, suggesting that a complex interplay between genetic and non-genetic factors affects weight.[3,4]

Advances in genotyping technologies have enabled the investigation of gene-environment (GxE) interactions.[4,5] For obesity outcomes, the ‘environment’ in GxE studies is often operationalised as the lifestyle or behavioural factors that influence energy balance,[6] rather than more upstream features of the built and social environments; the settings where behavioural ‘choices’ are made and constrained. Some recent studies have examined interactions between genetic risk and birth cohort as a means of capturing exposure to an increasingly obesogenic environment in very broad terms,[7–9] but there has been limited investigation of specific features of the environment that might plausibly interact with genetic risk,[10–14] despite a number of ‘socio-ecological’ environmental factors long being recognised in social epidemiology as potentially important determinants of weight status.

The residential neighbourhood environment comprises many features that potentially influence energy balance. These include the proximity, density and relative proportions of healthy and unhealthy food retailers,[15–17] and resources for physical activity (PA), such as leisure centres, swimming pools, gyms and sports fields.[18–21] Other neighbourhood features linked to energy balance include walkability, access to public transport and local resources such as public parks and greenspace.[22,23] If the genetic risk of obesity modifies the influence of these neighbourhood exposures, we would expect to observe differential effects of the residential environment on body mass index (BMI) according to level of genetic risk. The influence of the environment may be strongest in people with high genetic risk due to increased sensitivity to external factors,[24,25] or it may be strongest in people with low genetic risk, who maximise their genetic ‘advantage’ within a healthier environment while those at greater risk express a higher BMI phenotype regardless of environmental factors.[6]

In this study we use the UK Biobank cohort to examine whether genetic risk of obesity modifies the effect of two residential environment exposures likely to influence BMI: proximity to fast-food and availability of formal PA facilities. We operationalise genetic risk in two ways. First, using polygenic risk scores derived from single nucleotide polymorphisms (SNPs) linked to BMI, and second, using the individual SNPs most strongly linked to BMI and thought to be involved in diet or PA pathways.

## METHODS

### Data

We used baseline data from UK Biobank.[26] Data were potentially available from 502,656 individuals who visited 22 UK Biobank assessment centres across the UK between 2006 and 2010. Individuals aged 40–69 years living within 25 miles of an assessment centre and listed on National Health Service (NHS) patient registers were invited to participate.

Linked to UK Biobank is the UK Biobank Urban Morphometric Platform (UKBUMP), a high-resolution spatial database of objectively measured characteristics of the physical environment surrounding each participant’s residential address, derived from multiple national spatial datasets.[27] Environmental measures include densities of various land uses and proximity to various health-relevant resources. Measures for the current study are available for 96% of the UK Biobank sample.

Genome-wide genetic data are available for 488,363 participants. Genetic data are missing from 3% of the sample as insufficient DNA was extracted from blood samples for genotyping assays. SNP genotypes not directly assayed were imputed. Procedures used to derive the genetic data and undertake quality assurance are reported in Bycroft et al.[28] Genetic data for the relevant SNPs were downloaded, decrypted and linked to participant IDs to facilitate analysis.

### Outcome

Body Mass Index (BMI, kg/m^2^) was calculated from weight and height measurements collected by trained staff using standard procedures.[26] The variable was normally distributed and analysed as a continuous outcome variable.

### Neighbourhood exposures

We examined interactions between genetic risk and two neighbourhood characteristics likely to influence BMI: availability of formal PA facilities (number of indoor and outdoor sporting and leisure facilities within a one-kilometre street-network distance of an individual’s home) and fast-food proximity (distance in metres to nearest takeaway/fast-food outlet). Greater neighbourhood availability of PA facilities may influence BMI through increased opportunities for physical activity, and greater distances from home to fast-food outlets may influence BMI by reducing access to fast food.[29,30] In prior analyses we found both were associated with BMI in the expected direction – that is, living further from a fast-food outlet, or having more PA facilities near home, was associated with having a lower BMI.[18] Both exposures were analysed as continuous variables, with higher values of each (more facilities; greater distance to nearest fast-food outlet) representing lower exposure. Due to the positively skewed distribution of these variables, number of PA facilities was capped at 15 (<1% recoded from >15) and distance to nearest fast-food outlet was log transformed (base 10) such that regression coefficients were interpreted as the mean difference in BMI associated with a 10-fold increase in distance to nearest fast-food outlet e.g. 100 metres to one kilometre.

### Genetic risk scores and individual SNPs

A recent genome-wide association study (GWAS) identified 97 SNPs associated with BMI.[31] We constructed a genetic risk score (GRS) based on 91 of these SNPS, excluding six SNPs identified elsewhere[32] as being in linkage disequilibrium with other included SNPs (rs17001654, rs2075650 and rs9925964) or having pleiotropic effects (rs11030104, rs3888190, rs13107325), both of which may produce bias in associations between the genetic risk score and the outcome, and in interaction analyses.[33] We also constructed an alternative GRS, the same as one by used Tyrrell and colleagues[32] in a study of UK Biobank participants of White British ancestry, in which they tested interactions between genetic risk and behavioural exposures using a GRS derived from 69 of the SNPs identified in the recent GWAS. Their GRS excluded SNPs from secondary meta-analyses of studies of regional, sex-stratified or non-European-descent populations,[31] and one SNP (rs2033529) that was unavailable at the time of their study. Full lists of the SNPs included in each of the 91-SNP and 69-SNP risk scores are provided in Supplementary Table 1. The GRSs were constructed by summing the number of BMI-increasing alleles across the set of 69 or 91 loci, and weighting the allele count at each SNP by its published effect size.[31] For imputed SNP genotypes we used the imputed allelic dosages.

From the literature we identified individual SNPs with a well-established link to obesity and the largest published effect sizes (rs1558902 rs6567160 rs13021737, markers of the *FTO, MC4R* and *TMEM18* genes respectively),[1,31] and three SNPs recently linked to physical activity (rs13078960, rs10938397 and rs7141420, markers of *CADM2, GNPDA2 NRXN3*).[34,35] We tested for interactions between the number of BMI-increasing alleles at each of these loci, and each neighbourhood variable. We hypothesise that if GxE interactions are observed for these SNPs, those SNPs implicated in dietary behaviour will only interact with the fast-food environment, and those implicated in PA behaviour will only interact with the PA environment.

### Covariates

Models were adjusted for potential confounding by age, sex, educational attainment, household income, employment status, area deprivation (Townsend score), urban/non-urban status, and neighbourhood residential density and mutually adjusted for the other neighbourhood exposure. We also corrected for population stratification by adjusting for the first ten of 40 UK Biobank-provided genetic ancestry principal components from a genome-wide PCA of UK Biobank’s genetic data.[28]

### Statistical analysis & analytic sample

Accounting for the nested structure of the data (individuals within assessment areas), we used mixed effects models with a random coefficient for the neighbourhood exposure and assuming an unstructured variance/covariance matrix. Models included an interaction term between the neighbourhood exposure and the genetic risk score, with both analysed as continuous variables. BMI difference per unit change in the exposure was estimated for each quintile of genetic risk. The *p*-value for the additive interaction term was interpreted as strength of evidence of effect modification. The marginal predicted values of BMI associated with different levels of each neighbourhood exposure from these models were plotted for the top and bottom quintile of genetic risk, to visualise observed effect heterogeneity according to genetic risk. A complete case analysis was used, restricted to UK Biobank participants of White British ancestry (defined by concordant self-report and PCA results for White British/Caucasian ancestry) for the primary analyses because the smaller GRS was limited to SNPs associated with BMI in analyses of individuals with European ancestry. The sample size for the primary analysis was 335,046. Analysis was performed using Stata SE v14.2.

### Sensitivity analyses

As the 91-SNP GRS included SNPs associated with BMI in populations of non-European descent, we undertook a sensitivity analysis that tested for an interaction with the 91-SNP GRS in a sample unrestricted by ethnicity to test generalisability to the wider source population. To explore the possibility that results might be biased by latent genetic structure in the sample – a concern regarding genetic analyses involving UK Biobank[36] – we also performed sensitivity analyses in which models were adjusted for all 40 genetic ancestry principal components, and for birth location. Finally, although weighting of the polygenic risk scores is appropriate due to the varying degree to which each SNP is associated with BMI, we performed sensitivity analyses using an unweighted version of each GRS. Evidence of a GxE interaction using unweighted scores is expected to be weaker, due to dilution of the effects of the more influential SNPs.

### Ethics

UK Biobank has ethical approval from the North West Multi-centre Research Ethics Committee (reference 16/NW/0274), the Patient Information Advisory Group (PIAG), and the Community Health Index Advisory Group (CHIAG). Additional ethical approval for the specific study was obtained from the London School of Hygiene and Tropical Medicine’s Research Ethics Committee in September 2016 (reference 11897).

## RESULTS

The sample was 52.2% female, with a mean age of 56.5 years (range 40-70 years at baseline). Mean BMI was 27.4 kg/m^2^ (SD=4.7), median distance to nearest fast-food outlet was 1171 metres and median number of PA facilities within one kilometre of home was one. Sample characteristics are summarised in Table 1.

**Table 1.** Characteristics of total sample and top and bottom quintile of 91-SNP genetic risk score

|  | 91-SNP genetic risk score |  | Total sample |
| --- | --- | --- | --- |
|  | Quintile 1<br>(lowest risk of<br>obesity) | Quintile 5<br>(highest risk of<br>obesity) |  |
| Total number of participants | 64269 | 69577 | 335046 |
| BMI (kg/m<sup>2</sup>) (*mean, SD*) | 26.5 (4.3) | 28.3 (5.1) | 27.4 (4.7) |
| Distance to nearest fast-food outlet<br>(m) (*median, IQR*) | 1172 (634 - 2302) | 1169 (626 - 2290) | 1171 (630-2301) |
| Number of PA facilities within 1km of<br>home address (*median, IQR*) | 1 (0 - 3) | 1 (0 - 3) | 1 (0 - 3) |
| Age (*mean, SD*) | 56.5 (8.0) | 56.5 (8.0) | 56.5 (8.0) |
| Sex (female) | 33876 (52.7%) | 35923 (51.6%) | 174872 (52.2%) |
| Income |  |  |  |
| Less than 18,000 | 14154 (22.0%) | 15734 (22.6%) | 74556 (22.3%) |
| 18,000 to 30,999 | 16497 (25.7%) | 18270 (26.3%) | 86917 (25.9%) |
| 31,000 to 51,999 | 17013 (26.5%) | 18374 (26.4%) | 88721 (26.5%) |
| 52,000 to 100,000 | 13269 (20.7%) | 13738 (19.8%) | 67908 (20.3%) |
| Greater than 100,000 | 3336 (5.2%) | 3461 (5.0%) | 16944 (5.1%) |
| Education |  |  |  |
| College or University degree | 21462 (33.4%) | 22412 (32.2%) | 110153 (32.9%) |
| A levels/AS levels or equivalent | 7635 (11.9%) | 7961 (11.4%) | 39017 (11.7%) |
| O levels/GCSEs or equivalent | 14262 (22.2%) | 15779 (22.7%) | 74966 (22.4%) |
| CSEs or equivalent | 3500 (5.5%) | 3933 (5.7%) | 18722 (5.6%) |
| NVQ or HND or HNC or equivalent | 4266 (6.6%) | 4782 (6.9%) | 22892 (6.8%) |
| Other professional qualifications | 3302 (5.1%) | 3527 (5.1%) | 16954 (5.1%) |
| None of the above | 9842 (15.3%) | 11183 (16.1%) | 52342 (15.6%) |
| Employment status |  |  |  |
| Paid employment or self-employed | 38217 (59.5%) | 41326 (59.4%) | 199280 (59.5%) |
| Retired | 21330 (33.2%) | 23096 (33.2%) | 111113 (33.2%) |
| Unable to work | 1756 (2.7%) | 2055 (3.0%) | 9457 (2.8%) |
| Unemployed | 769 (1.2%) | 878 (1.3%) | 4238 (1.3%) |
| Home duties/carers/student/other | 2197 (3.4%) | 2222 (3.2%) | 10958 (3.3%) |
| Urbanicity (% urban dwelling) | 54560 (84.9%) | 59018 (84.8%) | 284471 (84.9%) |
| Area deprivation<sup>a</sup> (*mean, SD*) | -1.6 (2.9) | -1.6 (2.9) | -1.6 (2.9) |
| Residential density<sup>b</sup> (*median, IQR*) | 1794 (1041 - 2934) | 1801 (1043 - 2911) | 1798 (1044 - 2918) |

<sup>a</sup> 2001 Townsend index score<sup>b</sup> Residential address points per 1km street-network buffer

Using the two alternative weighted genetic risk scores, we observed evidence of an interaction between fast-food proximity and genetic risk (P=0.028 for the 91-SNP GRS, P=0.018 for the 69-SNP GRS). The magnitude of the estimated effect between fast-food proximity and BMI was small at all levels of genetic risk, but increased as genetic risk increased. In the highest quintile of genetic risk of obesity (based on the 91-SNP GRS), each 10-fold increase in distance to the nearest fast-food store was associated with a 0.194kg/m^2^ lower mean BMI (95%CI: -0.326,-0.062), which was twice the magnitude of association in the lowest risk quintile (β=-0.081; 95%CI: -0.213,0.052) (Table 2; Figure 1).

**Table 2.** Associations between neighbourhood variables and BMI, by quintile of genetic risk based on 91-SNP and 69-SNP risk scores

|  | 91-SNP GRS |  |  | 69-SNP GRS |  |  |
| --- | --- | --- | --- | --- | --- | --- |
|  | Quintile of genetic risk | Mean BMI difference for unit increase in neighbourhood exposure | P-interaction | Quintile of genetic risk | Mean BMI difference for unit increase in neighbourhood exposure | P-interaction |
| **Fast-food proximity<sup>a,b</sup>**<br>(beta represents BMI difference for a 10-fold increase in distance (m) to nearest fast-food outlet) | Q1 | -0.081 (-0.213, 0.052) | 0.028 | Q1 | -0.080 (-0.214, 0.055) | 0.018 |
|  | Q2 | -0.115 (-0.239, 0.009) |  | Q2 | -0.117 (-0.243, 0.009) |  |
|  | Q3 | -0.137 (-0.259, -0.014) |  | Q3 | -0.140 (-0.264, -0.017) |  |
|  | Q4 | -0.158 (-0.282, -0.035) |  | Q4 | -0.164 (-0.289, -0.039) |  |
|  | Q5 | -0.194 (-0.326, -0.062) |  | Q5 | -0.204 (-0.337, -0.070) |  |
| **Availability of PA facilities<sup>a,c</sup>**<br>(beta represents BMI difference for each additional facility) | Q1 | -0.074 (-0.100, -0.047) | 0.530 | Q1 | -0.070 (-0.097, -0.044) | 0.178 |
|  | Q2 | -0.075 (-0.101, -0.049) |  | Q2 | -0.074 (-0.099, -0.048) |  |
|  | Q3 | -0.076 (-0.101, -0.050) |  | Q3 | -0.076 (-0.101, -0.050) |  |
|  | Q4 | -0.077 (-0.103, -0.051) |  | Q4 | -0.078 (-0.103, -0.052) |  |
|  | Q5 | -0.078 (-0.105, -0.052) |  | Q5 | -0.081 (-0.106, -0.054) |  |

<sup>a</sup> Regression models were adjusted for age (years), sex (male/female), highest education level attained (Degree; A level or equivalent; O level or equivalent; CSE or equivalent; NVQ/HND/HNC; other professional qualification; none of the above), annual household income (<£18,000; £18,000-£30,999; £31,000-£51,999; £52,000-£100,000; >£100,000), employment status (paid work, retired, unable to work, unemployed, other), area deprivation (Townsend score), urbanicity (urban/non-urban), neighbourhood residential density (count of residential features within a one-km street network buffer of home address, log transformed).
<sup>b</sup> Also adjusted for availability of PA facilities.
<sup>c</sup> Also adjusted for fast-food proximity.

**Figure 1.**
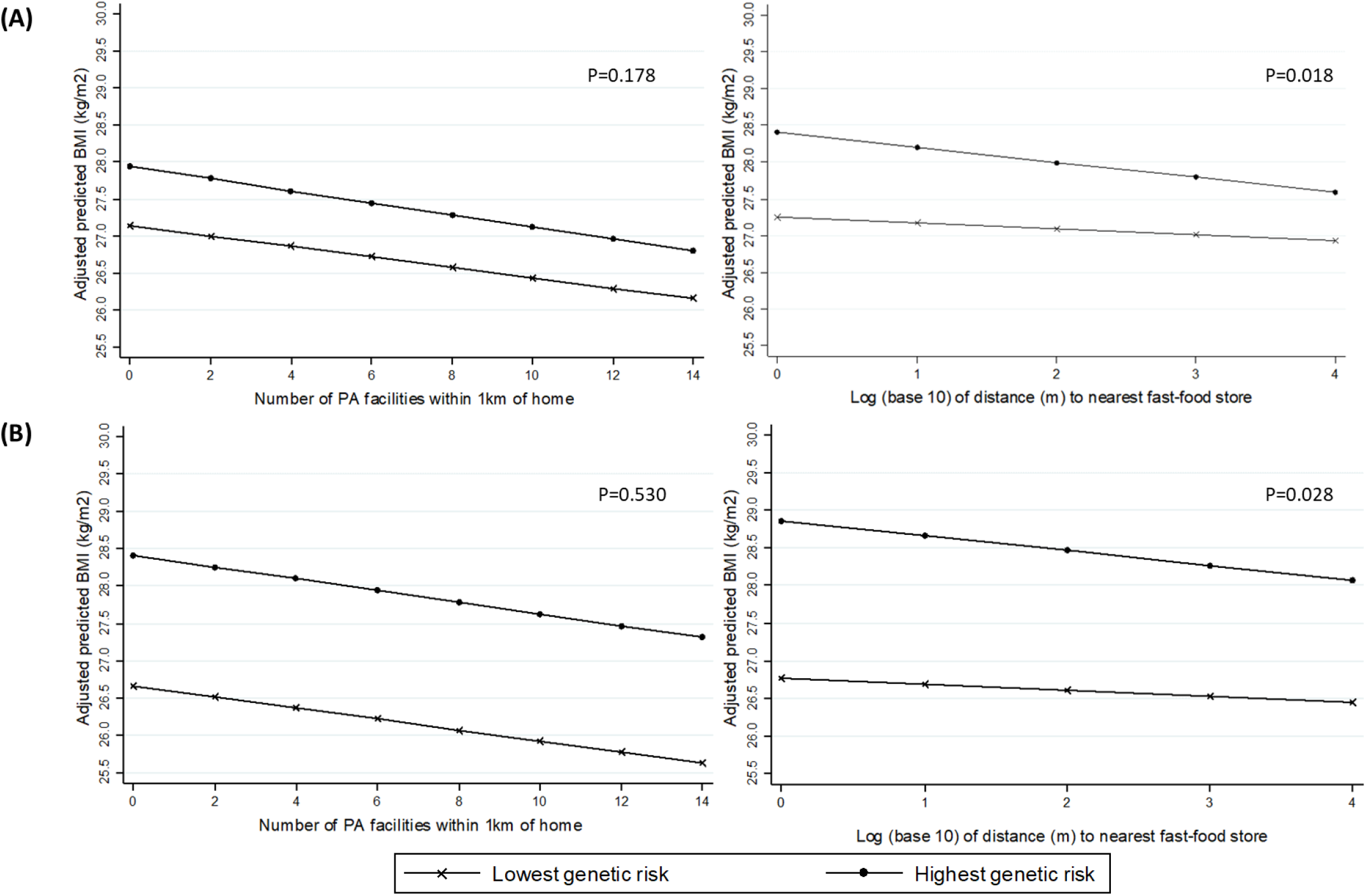
Association between neighbourhood variables and BMI in the highest and lowest quintiles of genetic risk, based on (a) 69-SNP Genetic Risk Score, and (b) 91-SNP Genetic Risk Score

There was less evidence that the association between availability of PA facilities and BMI was modified by genetic risk. The magnitude of the association between number of formal PA facilities within 1km of home and BMI was similar at all levels of genetic risk, and while effect estimates did increase slightly with increasing genetic risk, differences between risk groups were small with weak evidence of interaction for both the 91-SNP GRS (P=0.530) and 69-SNP GRS (P=0.178). For both environmental exposures, the results obtained from the two different weighted GRSs were substantively similar, but evidence of an interaction with the 69-SNP GRS was somewhat stronger (Table 2; Figure 1). The plots in Figure 1 also demonstrate that the BMI difference between the highest and lowest risk quintiles is greater for the 91-SNP GRS than the 69-SNP GRS, reflecting the fact that the larger GRSs captures more of the genetic variation in BMI.

Examination of interactions between neighbourhood variables and specific SNPs revealed strong evidence of one interaction: with the marker of *MC4R*, which encodes the melanocortin-4 receptor previously shown to be important in the regulation of food intake. Among people homozygous for the high risk allele at the marker of *MC4R*, each 10-fold increase in distance to the nearest fast-food store was associated with a 0.258 kg/m^2^ lower BMI, compared with only a 0.096 kg/m^2^ difference per 10-fold increase in distance among people with no risk alleles at this locus ((P_interaction_=0.009, Table 3; Figure 2). Some evidence of an interaction between fast-food proximity and rs1558902, the marker of the *FTO* gene (P=0.067), where again the higher risk group showed a stronger association between fast-food proximity and BMI. We also observed some evidence of a GxE interaction between the availability of PA facilities and rs13021737 (in the *TMEM18* gene) (P=0.076). In this case, increased genetic risk attenuated the association between availability of PA facilities and BMI (Figure 2).

**Table 3.** Association between neighbourhood variables and BMI, testing interaction with number of risk alleles at selected loci

| **rs1558902 (*FTO*)** |  |  |  |  |
| --- | --- | --- | --- | --- |
|  | P-interaction | Homozygous low risk<br>(0 risk alleles) | Heterozygous<br>(1 risk allele) | Homozygous high risk<br>(2 risk alleles) |
| **Fast-food proximity** | 0.067 | -0.099 (-0.198, -0.001) | -0.148 (-0.238, -0.059) | -0.197 (-0.305, -0.088) |
| **Availability of PA facilities** | 0.933 | -0.077 (-0.104, -0.050) | -0.077 (-0.103, -0.051) | -0.076 (-0.104, -0.049) |
| **rs6567160 (*MC4R*)** |  |  |  |  |
|  | P-interaction | Homozygous low risk<br>(0 risk alleles) | Heterozygous<br>(1 risk allele) | Homozygous high risk<br>(2 risk alleles) |
| **Fast-food proximity** | 0.009 | -0.096 (-0.188, -0.003) | -0.177 (-0.271, -0.083) | -0.258 (-0.386, -0.130) |
| **Availability of PA facilities** | 0.606 | -0.078 (-0.104, -0.051) | -0.075 (-0.102, -0.049) | -0.073 (-0.103, -0.043) |
| **rs13021737 (*TMEM18*)** |  |  |  |  |
|  | P-interaction | Homozygous low risk<br>(0 risk alleles) | Heterozygous<br>(1 risk allele) | Homozygous high risk<br>(2 risk alleles) |
| **Fast-food proximity** | 0.993 | -0.135 (-0.226, -0.043) | -0.135 (-0.234, -0.036) | -0.135 (-0.279, 0.008) |
| **Availability of PA facilities** | 0.076 | -0.080 (-0.106, -0.053) | -0.071 (-0.098, -0.043) | -0.061 (-0.093, -0.030) |
| **rs13078960 (*CADM2*)** |  |  |  |  |
|  | P-interaction | Homozygous low risk<br>(0 risk alleles) | Heterozygous<br>(1 risk allele) | Homozygous high risk<br>(2 risk alleles) |
| **Fast-food proximity** | 0.114 | -0.159 (-0.252, -0.066) | -0.108 (-0.205, -0.010) | -0.056 (-0.192, 0.081) |
| **Availability of PA facilities** | 0.419 | -0.076 (-0.102, -0.049) | -0.079 (-0.106, -0.053) | -0.083 (-0.114, -0.053) |
| **rs10938397 (*GNPDA2*)** |  |  |  |  |
|  | P-interaction | Homozygous low risk<br>(0 risk alleles) | Heterozygous<br>(1 risk allele) | Homozygous high risk<br>(2 risk alleles) |
| **Fast-food proximity** | 0.328 | -0.115 (-0.215, -0.015) | -0.141 (-0.230, -0.052) | -0.167 (-0.274, -0.061) |
| **Availability of PA facilities** | 0.694 | -0.076 (-0.102, -0.049) | -0.077 (-0.103, -0.051) | -0.079 (-0.106, -0.051) |
| **rs7141420 (*NRXN3*)** |  |  |  |  |
|  | P-interaction | Homozygous low risk<br>(0 risk alleles) | Heterozygous<br>(1 risk allele) | Homozygous high risk<br>(2 risk alleles) |
| **Fast-food proximity** | 0.520 | -0.152 (-0.257, -0.048) | -0.135 (-0.227, -0.043) | -0.118 (-0.224, -0.012) |
| **Availability of PA facilities** | 0.125 | -0.071 (-0.097, -0.044) | -0.077 (-0.102, -0.051) | -0.083 (-0.110, -0.056) |

**Figure 2.**
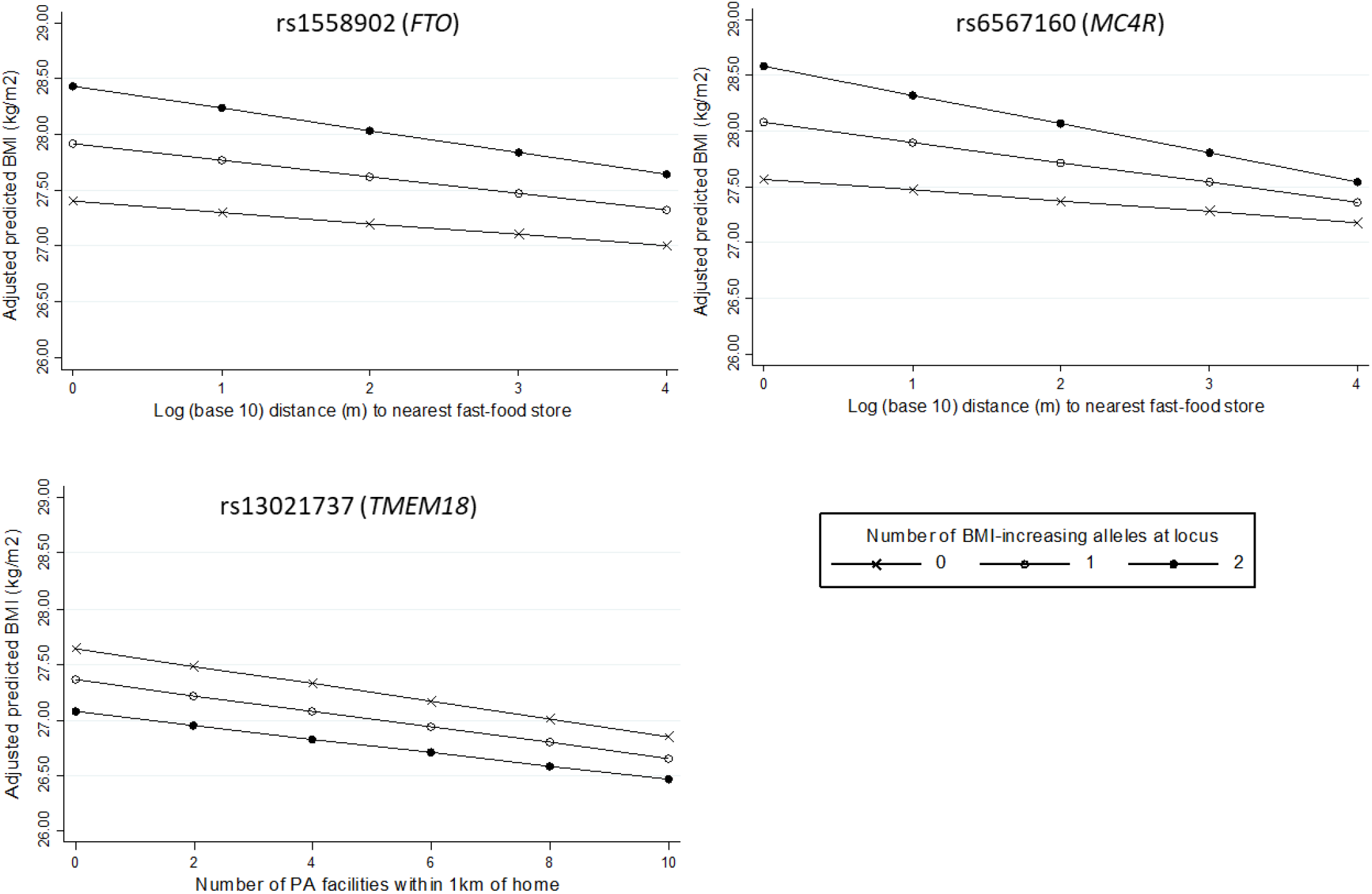
Association between neighbourhood variables and BMI according to number of risk alleles at individual SNPs where P_interaction_<0.10 (rs1558902 & rs6567160 for fast-food proximity; rs13021737 for availability of PA facilities)

In sensitivity analyses, interactions between fast-food proximity and genetic risk were – as expected – weaker when the genetic risk scores were not weighted by the effect sizes of the component SNPs, with mean differences in BMI more similar across levels of genetic risk than we observed using the weighted score (Supplementary Table 2). Expanding the sample to include non-White ethnicities, we observed slightly increased P-values for the interaction terms but otherwise no substantive difference from the primary analysis (Supplementary Table 3). For all models, the impact of adjusting for 40 rather than 10 genetic ancestry principal components was negligible, while some attenuation of the interaction between fast-food proximity and polygenic risk occurred when adjusting for birth location (Supplementary Table 4).

## DISCUSSION

In UK Biobank we found evidence that genetic risk of obesity modifies sensitivity to the neighbourhood food environment, though effects are small. We found that people at higher genetic risk of obesity have higher average BMI the closer they live to a fast-food outlet, whereas for those at low genetic risk of obesity, distance to the nearest fast-food outlet does not appear to be associated with BMI. In contrast, an overall negative association between neighbourhood availability of PA facilities and BMI varies very little across levels of polygenic risk.

The observed gene-environment interaction for fast-food proximity using polygenic risk scores was supported by stronger evidence of an interaction between fast-food proximity and a specific SNP near *MC4R*, a gene known to be involved in regulation of food intake[37]. Previous research has linked *MC4R* specifically to binge eating[38] although this remains contested.[39] We also observed some evidence of a possible interaction with a SNP marker of *FTO*, a gene with well-established links to obesity. While *FTO* has long been recognised as an obesity-associated locus, and has been implicated in central nervous system regulation of appetite, its exact function remains poorly understood.[1] In a study of gene-diet interactions, genetic risk scores for BMI were found to be associated with fried food consumption, and, consistent with our results, individual loci in or near both *MC4R* and *FTO* contributed to this.[40]

Limited evidence for an interaction between genetic risk and the PA environment is consistent with findings from a recent study in adolescents that found that availability of recreation facilities did not contribute to the attenuation by PA of genetic risk of obesity.[34] While overall genetic risk of obesity did not interact with the PA environment in our study, the weaker association we observed between the availability of PA facilities and BMI in those with more risk alleles at the *TMEM18* locus suggests that some specific SNPs might. Further examination of other SNPs is warranted. Lack of interaction with specific SNPs might be explained by the pathways they influence being less sensitive to environmental exposures. As the functional pathways by which most BMI-associated loci influence BMI remain poorly understood, it is difficult to speculate further.

Stronger evidence for interactions with specific SNPs highlights the lack of specificity of polygenic risk scores. While useful in exploratory studies, grouping all SNPs statistically associated with a complex phenotype such as BMI into a single score, regardless of the function of the genes they represent, may dilute or obscure important interactions. Scores based on known or putative biological mechanisms may prove more valuable, particularly for elucidating causal relationships. We observed very similar results for both the 69-SNP and 91-SNP genetic risk scores, although the smaller GRS yielded stronger evidence of interaction. It may be that the additional SNPs in the larger GRS diluted the interaction due to being associated with BMI only in some population subgroups, and some having been linked to BMI only in more ethnically diverse populations than our primary sample.

We have reported elsewhere that the main association between fast-food proximity and BMI in UK Biobank may be attenuated due to measurement error in the exposure,[18] and because the exposure does not account for other, healthier elements of the food environment.[41] Compared with other measures of the fast-food environment, proximity measures may also produce more conservative estimates of association with relevant outcomes.[42] In a regional sub-sample of UK Biobank, others have recently improved on this measurement of the food environment and found stronger associations.[43] In this study, where the main effect sizes are relatively small, even the reasonably strong interaction effects we observed translate to small differences between high and low risk groups. However, given the likely measurement error and the distal and complex nature of the relationships under investigation, detecting even weak associations and small differences might point to potentially important processes. Other studies have reported evidence of a GxE interaction between genetic risk of obesity and birth cohort[7–9], with this interpreted as evidence that recent increases in the ‘obesogenicity’ of our environments increases the susceptibility of those with a genetic predisposition to become obese. Here we examined two characteristics of neighbourhood environments likely to be obesogenic; but others may also interact with genetic risk. For example, GxE interactions have recently been reported for neighbourhood walkability and obesity,[13] and neighbourhood deprivation and BMI.[12] Given that unhealthy characteristics of neighbourhoods often cluster together,[44] the combined effects of multiple ‘obesogenic’ features on those at increased genetic risk of obesity may be substantial.

Our findings provide evidence for a potentially important GxE interaction, but further confirmatory studies are required. Another recent study found a strong GxE interaction between genetic risk of obesity and socioeconomic status, and while our analyses are adjusted for several socioeconomic indicators, if there remains any residual confounding by SES it may be contributing to the GxE interactions we observed. Geographical genetic structure in the sample remains a risk, even after adjustment for ancestry components and geography. Such structure may induce spurious associations with polygenic risk scores in particular.[36] In sensitivity analyses we found that adjustment for additional ancestry principal components had negligible impact on the strength of evidence for the GxE interactions we tested, but evidence for a genetic interaction with fast-food proximity was slightly weaker following adjustment for birth location. Further investigation of the effect of the residual genetic structure in the sample is warranted. GxE interactions are also sensitive to the scaling of environmental variables, and the power to detect a GxE interaction can depend on the main effect sizes, and distribution and measurement quality of the genetic and environmental variables.[45] Studies using UK Biobank are also at risk of selection bias due to a low response rate.[46] It is important these analyses are replicated in other samples at lower risk of these biases.

It is widely accepted that environmental factors are important in explaining the recent rise in the global prevalence of overweight and obesity. In this study, we find evidence that people at higher genetic risk of obesity may more sensitive to exposure to the residential fast-food environment. Ensuring that neighbourhood residential environments are designed to promote a healthy weight may be particularly important for those with genetic susceptibility to obesity.

## Data Availability

Data are available from UK Biobank to approved researchers

https://www.ukbiobank.ac.uk/

## ACKNOWLEDGEMENTS

This research was conducted using the UK Biobank Resource under Application Number 17380. The authors wish to acknowledge the work of the UK Biobank research team, including those who generated the UK Biobank Urban Morphometric Platform, and thank all the UK Biobank participants. KM was funded by a Commonwealth Scholarship Commission PhD Scholarship. NP was supported by the Wellcome Trust Institutional Strategic Support Fund, 097834/Z/11/B through the Centre for Global NCDs and SC is supported by Health Data Research UK. Our funders had no role in any stage of this study, nor in the preparation of the manuscript for publication.

## Author contributions

SC and KM conceived of the study, and KM, LP and SC designed the analysis. KM led the data management, statistical analysis, and writing of the manuscript. JP extracted the genetic data from UK Biobank, and JP and LP provided expertise in working with genetic data. SC, LP and NP contributed to the interpretation of results, and all authors contributed to and approved of the final manuscript.

## REFERENCES

1 Waalen J. The genetics of human obesity. Transl Res 2014;164:293–301. doi:10.1016/j.trsl.2014.05.010

2 Popkin BM, Gordon-Larsen P. The nutrition transition: worldwide obesity dynamics and their determinants. Int J Obes Relat Metab Disord 2004; 28 Suppl 3:S2–9. doi:10.1038/sj.ijo.0802804

3 Van Der Klaauw AA, Farooqi IS. The hunger genes: Pathways to obesity. Cell 2015;161:119–32. doi:10.1016/j.cell.2015.03.008

4 Richmond RC, Timpson NJ. Recent Findings on the Genetics of Obesity: Is there Public Health Relevance? Curr Nutr Rep 2012;1:239–48. doi:10.1007/s13668-012-0027-x

5 Andreasen CH, Andersen G. Gene-environment interactions and obesity--further aspects of genomewide association studies. Nutrition 2009;25:998–1003. doi:10.1016/j.nut.2009.06.001

6 Boardman JD, Daw J, Freese J. Defining the environment in gene-environment research: Lessons from social epidemiology. Am J Public Health 2013;103:S64–72. doi:10.2105/AJPH.2013.301355

7 Rosenquist JN, Lehrer SF, O’Malley AJ, et al. Cohort of birth modifies the association between FTO genotype and BMI. Proc Natl Acad Sci U S A 2015;112:354–9. doi:10.1073/pnas.1411893111

8 Walter S, Mejía-Guevara I, Estrada K, et al. Association of a Genetic Risk Score With Body Mass Index Across Different Birth Cohorts. JAMA 2016;316:63. doi:10.1001/jama.2016.8729

9 Brandkvist M, Bjørngaard JH, Ødegård RA, et al. Quantifying the impact of genes on body mass index during the obesity epidemic: longitudinal findings from the HUNT Study. BMJ 2019;366:1–8. doi:10.1136/bmj.l4067

10 Horn EE, Turkheimer E, Strachan E, et al. Behavioral and Environmental Modification of the Genetic Influence on Body Mass Index: A Twin Study. Behav Genet 2015;45:409–26. doi:10.1007/s10519-015-9718-6

11 Mooney SJ, Grady ST, Sotoodehnia N, et al. In the wrong place with the wrong SNP. Epidemiology 2016;27:656–62. doi:10.1097/EDE.0000000000000503

12 Owen G, Jones K, Harris R. Does neighbourhood deprivation affect the genetic influence on body mass? Soc Sci Med 2017;185:38–45. doi:10.1016/j.socscimed.2017.05.041

13 Kowaleski-Jones L, Brown BB, Fan JX, et al. The joint effects of family risk of obesity and neighborhood environment on obesity among women. Soc Sci Med 2017;195:17–24. doi:10.1016/j.socscimed.2017.10.018

14 Robinette JW, Boardman JD, Crimmins EM. Differential vulnerability to neighbourhood disorder: A gene×environment interaction study. J Epidemiol Community Health 2019;73:388–92. doi:10.1136/jech-2018-211373

15 Fleischhacker SE, Evenson KR, Rodriguez DA, et al. A systematic review of fast food access studies. Obes Rev 2011;12:e460–71. doi:10.1111/j.1467-789X.2010.00715.x

16 Burgoine T, Forouhi NG, Griffin SJ, et al. Associations between exposure to takeaway food outlets, takeaway food consumption, and body weight in Cambridgeshire, UK: Population based, cross sectional study. BMJ 2014;348:g1464. doi:10.1136/bmj.g1464

17 Black C, Moon G, Baird J. Dietary inequalities: what is the evidence for the effect of the neighbourhood food environment? Health Place 2014;27:229–42. doi:10.1016/j.healthplace.2013.09.015

18 Mason KE, Pearce N, Cummins S. Associations between fast food and physical activity environments and adiposity in mid-life: cross-sectional, observational evidence from UK Biobank. Lancet Public Heal 2018;3:e24–33. doi:10.1016/S2468-2667(17)30212-8

19 Ellaway A, Lamb KE, Ferguson NS, et al. Associations between access to recreational physical activity facilities and body mass index in Scottish adults. BMC Public Health 2016;16:756. doi:10.1186/s12889-016-3444-8

20 Van Holle V, Deforche B, Van Cauwenberg J, et al. Relationship between the physical environment and different domains of physical activity in European adults: a systematic review. BMC Public Health 2012;12:807. doi:10.1186/1471-2458-12-807

21 Barrientos-Gutierrez T, Moore KAB, Auchincloss AH, et al. Neighborhood Physical Environment and Changes in Body Mass Index: Results From the Multi-Ethnic Study of Atherosclerosis. Am J Epidemiol 2017;186:1–9. doi:10.1093/aje/kwx186

22 Townshend T, Lake AA. Obesogenic urban form: theory, policy and practice. Health Place 2009;15:909–16. doi:10.1016/j.healthplace.2008.12.002

23 Black JL, Macinko J. Neighborhoods and obesity. Nutr Rev 2008;66:2–20.

24 Davis OSP, Haworth CMA, Lewis CM, et al. Visual analysis of geocoded twin data puts nature and nurture on the map. Mol Psychiatry 2012;17:867–74. doi:10.1038/mp.2012.68

25 Llewellyn C, Wardle J. Behavioral susceptibility to obesity: Gene-environment interplay in the development of weight. Physiol Behav 2015;152:494–501. doi:10.1016/j.physbeh.2015.07.006

26 UK Biobank. UK Biobank: Protocol for a large-scale prospective epidemiological resource. 2007.

27 Sarkar C, Webster C, Gallacher J. UK Biobank Urban Morphometric Platform (UKBUMP) – A nationwide resource for evidence-based healthy city planning and public health interventions. Ann GIS 2015;21:135–48. doi:10.1080/19475683.2015.1027791

28 Bycroft C, Freeman C, Petkova D, et al. The UK Biobank resource with deep phenotyping and genomic data. Nature 2018;562:203–9. doi:10.1038/s41586-018-0579-z

29 Block J, Seward M, James P. Food environment and health. In: Duncan DT, Kawachi I, eds. Neighborhoods and Health. New York: : Oxford University Press 2018. 247–77.

30 Sallis JF, Glanz K. Physical Activity and Food Environments: Solutions to the Obesity Epidemic. Milbank Q 2009;87:123–54. doi:10.2307/25474362

31 Locke AE, Kahali B, Berndt SI, et al. Genetic studies of body mass index yield new insights for obesity biology. Nature 2015;518:197–206. doi:10.1038/nature14177

32 Tyrrell J, Wood AR, Ames RM, et al. Gene-obesogenic environment interactions in the UK Biobank study. Int J Epidemiol 2017;46:559–75. doi:10.1093/ije/dyw337

33 Dudbridge F, Fletcher O. Gene-environment dependence creates spurious gene-environment interaction. Am J Hum Genet 2014;95:301–7. doi:10.1016/j.ajhg.2014.07.014

34 Graff M, Richardson AS, Young KL, et al. The interaction between physical activity and obesity gene variants in association with BMI: Does the obesogenic environment matter? Health Place 2016;42:159–65. doi:10.1016/j.healthplace.2016.09.003

35 Klimentidis YC, Raichlen DA, Bea J, et al. Genome-wide association study of habitual physical activity in over 377,000 UK Biobank participants identifies multiple variants including CADM2 and APOE. Int J Obes 2018;42:1161–76. doi:10.1038/s41366-018-0120-3

36 Haworth S, Mitchell R, Corbin L, et al. Apparent latent structure within the UK Biobank sample has implications for epidemiological analysis. Nat Commun 2019;10:333. doi:10.1038/s41467-018-08219-1

37 Spiegelman BM, Flier JS. Obesity and the regulation of energy balance. Cell 2001;104:531–43. doi:10.1016/S0092-8674(01)00240-9

38 Branson R, Potoczna N, Kral JG, et al. Binge Eating as a Major Phenotype of Melanocortin 4 Receptor Gene Mutations. N Engl J Med 2003;348:1096–103. doi:10.1056/NEJMoa021971

39 Valette M, Bellisle F, Carette C, et al. Eating behaviour in obese patients with melanocortin-4 receptor mutations: A literature review. Int J Obes 2013;37:1027–35. doi:10.1038/ijo.2012.169

40 Qi Q, Chu AY, Kang JH, et al. Fried food consumption, genetic risk, and body mass index: gene-diet interaction analysis in three US cohort studies. BMJ 2014;348:g1610. doi:10.1136/bmj.g1610

41 Cummins S, Clary C, Shareck M. Enduring challenges in estimating the effect of the food environment on obesity. Am J Clin Nutr 2017;106:445–6. doi:10.3945/ajcn.117.161547

42 Bivoltsis A, Cervigni E, Trapp G, et al. Food environments and dietary intakes among adults: Does the type of spatial exposure measurement matter? A systematic review. Int J Health Geogr 2018;17:1–20. doi:10.1186/s12942-018-0139-7

43 Burgoine T, Sarkar C, Webster CJ, et al. Examining the interaction of fast-food outlet exposure and income on diet and obesity: Evidence from 51,361 UK Biobank participants. Int J Behav Nutr Phys Act 2018;15:1–12. doi:10.1186/s12966-018-0699-8

44 Fraser LK, Edwards KL, Cade J, et al. The geography of fast food outlets: A review. Int J Environ Res Public Health 2010;7:2290–308. doi:10.3390/ijerph7052290

45 Manuck SB, McCaffery JM. Gene-Environment Interaction. Annu Rev Psychol 2014;65:41–70. doi:10.1146/annurev-psych-010213-115100

46 Munafò MR, Tilling K, Taylor AE, et al. Collider scope: when selection bias can substantially influence observed associations. Int J Epidemiol 2017;:1–10. doi:10.1093/ije/dyx206

